# Social response to early-stage government control measures of COVID-19 in Colombia: population survey, April 8-20 2020

**DOI:** 10.1101/2020.06.18.20135145

**Authors:** Juan Carlos Rivillas-García, Rocío Murad-Rivera, Sandra Marcela Sánchez, Danny Rivera-Montero, Mariana Calderón-Jaramillo, Lina Maria Castaño, Marta Royo

## Abstract

On Monday, March 16, 2020, the government of Colombia announced actions to control COVID-19. These recommendations directly affected the entire population and included: reducing physical contact; reducing mobility and cancelling unnecessary travel; working from home; cancelling mass events; a 14 day at-home isolation period for people who arrived from international trips, or in cases in which someone in the household experienced a high temperature and sustained coughing; as well as increasing the frequency of hand washing and the use of face masks on public transport.

In order to understand the public sentiment around these recommendations, Asociación Profamilia developed an online survey through SurveyMonkey®. The survey was completed by 3549 adult people in Colombia (+18 years) between April 8 and April 20, 2020. In this report, we present the results of the survey:

- 98% think that the COVID-19 pandemic is a serious problem in Colombia.
- 90% are concerned that someone in their family will get COVID-19.
- 88% are concerned that someone in their family may have an emergency and not receive medical care.
- 46% believe they will likely get infected under the current Colombian government measures.
- 92% of adults reported taking at least one of the following measures to protect themselves from the COVID-19 infection:
  - 85% of senior citizens (60 years and older) voluntarily isolated or complied with government’s mandatory isolation measures.
  - 82% cut down on their mobility (avoided going out, using public transportation, and traveling).
  - 79% of those showing symptoms voluntarily isolated themselves and complied with government isolation measures.
  - 78% increased the frequency in which they use disinfectants, alcohol, and sanitizing gel, and how often they wash their hands.
  - 73% refrained from going out to social events and crowded places.
  - 70% avoided kissing and shaking hands.
  - 69% immediately complied with the government’s mandatory preventive isolation measures.
  - 63% avoided using public transportation.
  - 46% went into self-isolation (voluntary quarantine) before the government decreed it.
  - 32% started working from home.
- This study reveals that there are at least three groups of people in the country who are responding to the pandemic and physical distancing measures in different forms: those who resist the situation (34%), those who suffer from it (26%), and those who accept it (40%).
  - In the group of people who are resisting 40% are men; 68% are under 39 years old; 40% have savings and one out ten reported mental health problems; and 64% supported the government measures.
  - In the group of people suffering from the pandemic, 73% are women, 64% are under 29 years old, 55% have an average family income over 2 million pesos COP (roughly 417 GBP), 61% have had some chronic disease or somebody in the family; 73% reported mental problems. This group had the higher support and adherence to the government measures (68%).
  - In the group of people who are adapting to the situation, 76% are women, 43% are over 49 years old, 36% have savings, and 63% have not had chronic illnesses and 73% reported mental problems. This group had the lower support and adherence to the government measures (63%).
- 69% complied with government’s mandatory preventive isolation measures. This percentage was lower (64%) among young adults (25-29 years old).
- Overall, 83% are complying with preventive isolation and physical distancing. 77% feel that complying with isolation contributes to stopping COVID-19.
- Hygiene practices such as hand washing (78%), avoiding kissing and hand-shaking (70%), as well as using face masks (69%) were perceived as more effective measures to prevent the spread of the virus compared to physical distancing measures like cancelling travelling (46%), avoiding contact with people with fever or respiratory symptoms (43%), and avoiding contact with people who have travelled in the last month (35%).
- 80% live in the five cities with the highest spread of COVID-19.
  - 50% are responsible for the care of a family member; 16% are women heads of household.
  - 68% mentioned do not have savings.
  - 25% did not work before COVID-19.
- The top three concerns among the respondents were that someone in their family may get infected with COVID-19 (79%), that someone in their family may have an emergency and not receive medical care. (74%) and the fate of the poorest and most vulnerable people (71%).
- During quarantine, 75% have experienced issues with their mental health: 54% felt nervous; 52% have felt tired for no reason; 46% felt restless and impatient, and 34% felt anger and rage; 20% have experienced a need for sexual and reproductive health in the last 21 days which has not been met. 17% are concerned about their children’s bad behaviour and domestic violence.
- 63% get informed about COVID-19 through official websites; 52% have found false information about COVID-19 and the pandemic. 40% have experienced or witnessed jokes about the spread of COVID-19, and 25% have experienced or witnessed discrimination against health care personnel.
- 38% believe that the National Government’s response to control the virus was clear and consistent and 29% believe that it acted in a timely and swift manner; 46% believe that the Local Government’s response was clear and consistent and 42% believe that it acted in a quick and timely manner.

## Introduction

COVID-19 has caused a profound impact on global health, healthcare services, and the economy. It poses the most critical threat to public health from a respiratory syndrome since the 1918 H1N1 influenza pandemic (1). In the current absence of effective vaccines and pharmacological treatments, governments are responding with Non-Pharmacological Interventions (NPIs) to support mitigation measures (slowing the spread of the epidemic) and suppression (reversing epidemic growth) of COVID-19 to low levels (2). Non-pharmacological measures include the following:

1. Isolation of symptomatic cases at home: people with symptoms (cough and fever) remain at home for 7 days after symptoms begin.
2. Home quarantine for all family members of those with symptoms of the disease: stay at home for 14 days after symptoms start.
3. General physical distancing - a comprehensive policy to reduce contact with all people outside the home, school, or workplace, as well as limited mobility.
4. Physical distancing for people over 70 years old: only for those people over 70 years of age who are at the highest risk of suffering severe illness.
5. Closing of schools and universities.
6. Proper and frequent hand washing.

Evidence has shown why the COVID-19 outbreak grows exponentially and what are the resulting scenarios when pharmacological measures are put in place (2); it also shows the experience of some countries that implemented interventions early and were successful in reducing the number of cases when measures remained in place for several weeks, achieving a lower demand of healthcare services and lower mortality. However, the spread of the disease reappears once the controls are lifted (2,3).

In Colombia, the social response to non-pharmacological interventions, measures which aim to reduce contact between the people and thus reduce transmission of the virus, is yet to be evaluated. The implemented measures in Colombia during COVID-19 offer a unique and limited opportunity to generate relevant and prompt evidence on population’ perception and behavioural changes during preventive isolation and physical distancing.

Vaccine development has been deemed the most urgent research priority for many countries with research and development capacity. However, there is consensus on the fact that social studies that explore the experiences of the general population, their risk perceptions, and early-stage behaviours are equally necessary and essential (4).

### General Objective

To analyse the social adult response to early-stage government control measures of COVID-19 in Colombia.

#### Study design and survey implementation

- Cross-sectional survey using sub-national data. The total sample size was 3549 adults (18 years and over) in ten cities in Colombia.
- Non-probabilistic survey that was conducted between the 8th and 20th of April, 2020.
- Survey was conducted online using SurveyMonkey and sent to emails using Profamilia’s database, partners, and social networks (Twitter and Facebook, WhatsApp).

## Methods

### Study design and sampling

Cross-sectional survey using data at the subnational level. The online survey was created on **SurveyMonkey®**. We used snowball sampling because it allowed us to increase the sample size when those initially selected invite others to participate. The total number of completed surveys for analysis was 3,549. We selected five cities with the highest spread of the SARS-COV-2 virus and five cities with the lowest spread, according to reports from the Ministry of Health and Social Protection dated March 26, 2020. The cities with a high spread of the virus are Bogota (Bogota and greater Bogota); Cali (Cali and Yumbo); Medellin (Medellin and Aburra Valley); Cartagena; and Barranquilla (Barranquilla and Soledad). The selected cities with the lowest virus spread are Leticia, Riohacha, San Jose del Guaviare, Quibdo, and Sincelejo; additionally, 336 people from other municipalities completed the survey, and 19 people did not respond to the question on place of residence.

**Table 1.** **Cities with a high and low virus spread, number of conducted surveys and number of cases diagnosed with COVID-19**.

| Cities with high virus spread | Number of Surveys Conducted | Confirmed Cases | Cities with low virus spread | Number of Surveys Conducted | Confirmed Cases |
| --- | --- | --- | --- | --- | --- |
| Bogota and greater Bogota | 1291 | 1883 | Riohacha | 119 | 2 |
| Cali and greater Cali | 414 | 775 | Sincelejo | 80 | 1 |
| Medellin and greater Medellin | 762 | 426 | Quibdo | 43 | 10 |
| Barranquilla and greater Barranquilla | 240 | 125 | Leticia | 48 | 12 |
| Cartagena | 150 | 204 | San Jose del Guaviare | 47 | 0 |
| - | - | - | Other municipalities | 336 | 19*** |
| Total | 2857 | - | Total | 673 | - |

\* High and low spread of the virus with as of March 26, 2020.
\*\* Number of diagnosed cases as of April 21, 2020.
\*\*\* Average number of reported cases in other municipalities (1,003 cases distributed among 52 municipalities), taking into account the same municipalities that were found in the survey, for more information see Table 2.

The survey in Colombia has four components: 1) socio-demographic characteristics, 2) responsibility for care and employment, 3) risk and health perceptions, and 4) behavioural changes and capacity for isolation.

The considered socio-demographic characteristics were age, gender, residential area, vulnerable groups, education level, ethnicity, marital status, city of residence, home ownership, type of health insurance, employment status, income, and savings. Perceptions on risk and health were measured by perceived susceptibility and severity. To establish susceptibility, people were asked about their perceived probability of being infected with COVID-19 and about their general, mental, and sexual and reproductive health status under Colombian government preventive measures. Severity was measured by asking about how severe they perceived symptoms to be.

Behavioural changes included perceived effectiveness and adoption of preventive behaviours (to protect oneself and others) to avoid infection and subsequent transmission, based on three categories: (1) hygiene practices (use of face masks, washing hands more frequently with soap and water, using hand sanitizer more often, disinfecting surfaces and cleaning the house, covering nose and mouth when sneezing or coughing). (2) Travel cancellation (avoiding travelling to affected countries and urban and rural areas within Colombia, regardless of having contracted the virus). And finally (3), physical distancing (avoid public transportation, social events, going out, crowded places, and contact with people who have a fever or respiratory symptoms as well as going to the hospital only when necessary).

Two questions in this survey measured willingness and the ability for preventive isolation and physical distancing: i) In the last 21 days, which of the following measures have you taken to protect yourself and others from COVID-19? These include not going to work, school, or other public places, avoiding public transportation or taxis, and self-isolation or voluntary quarantine. The second question was: (ii) What were the reasons that motivated this change in behaviour?

One of the advantages of conducting an online survey using **SurveyMonkey®** is that it was easy and quick to use and that we could take advantage of the opportunity this quarantine brings, where many households have increased their connectivity and virtual activities. Among the limitations is the sampling method bias, as people tend to distribute the survey to people of equal socio-demographic characteristics. Therefore, the sample may report on specific population groups. A second limitation is that the results obtained are not representative of the country.

The information was collected, filtered, processed, and analyzed by Profamilia’s Research Directorate, and the Project and Research Management team in Bogota-Colombia. The research protocol was approved by Profamilia’s Research Ethics Committee (CEIP 2020-05) on April 8, 2020.

## Results

Generally speaking, the survey was completed in higher percentages by women (69%), people under 45 years of age (77%), people with a family income under five million COP (69%) (apx. 1,277 US), and people with a university education - undergraduate - (62%). The results are presented as follows:

1. Personal response and adoption of virus control measures.
2. Self-isolation (voluntary quarantine).
3. Groups that react under quarantine
4. Social response and protective measures in Colombia, Hong Kong, and the UK.
5. Care and employment responsibility.
6. Perception of risk, and unmet sexual, reproductive and mental health needs during the pandemic.
7. Access to quality information.
8. Perception of government response.

### 1. Personal response and adoption of virus control measures

82% reduced mobility and 78% of people increased their hand-washing frequency and use of disinfectant products, alcohol, or sanitizing gel (Figure 1).

**Figure 1.**
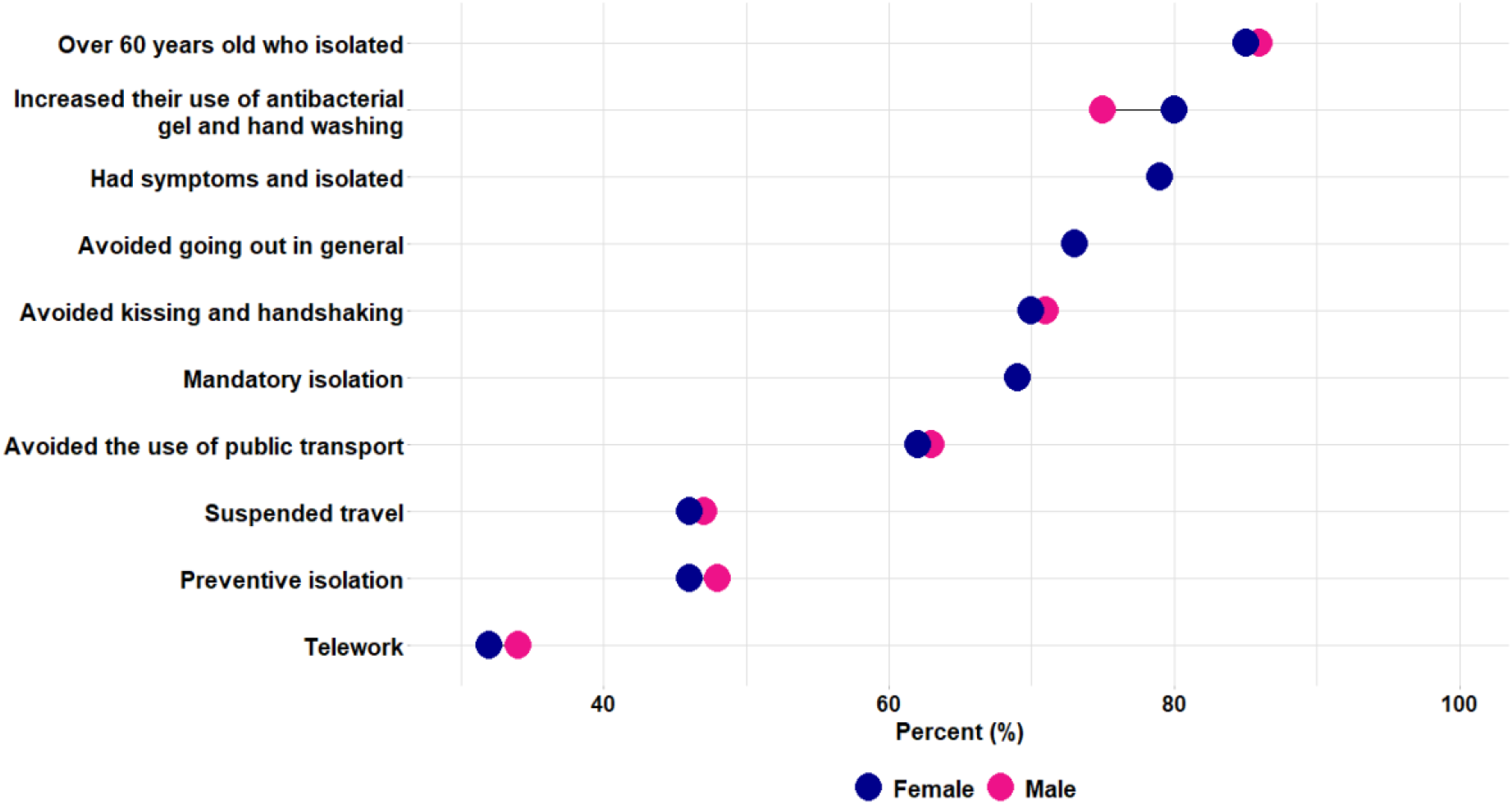
Percentage of respondents who adopted measures to protect themselves and others from COVID-19 by sex, Colombia 2020.

∘ 80% were women and 75% men.
∘ Those who used these measures over 80% were among 45-64-year-olds, while the lowest use (73%) was found among the youngest age group (18-24-year-olds).
∘ Cali and Bogotá (81%) have the highest levels of adoption, and the municipalities with the lowest virus spread (71%) present the lowest.

- 70% avoided kissing and shaking hands.
- 69% used face masks.
- 69% immediately complied with mandatory preventive isolation.
- 65% cover their mouth with their elbow when sneezing or coughing.
- 62% avoided public transportation.
- 59% increased the clean-up frequency of their home/workplace.
- 58% are more alert to their body’s symptoms.
- 54% avoid visiting healthcare centres and hospitals.
- 46% cancelled trips.
- 46% voluntarily went into isolation before the government decreed it.
- 43% avoided contact with people who had fever or respiratory symptoms.
- 35% avoided contact with people who had travelled in the last month.
- 12% avoided sex with their couple, spouse, sexual partner, significant other.
- 0.3% took no action to protect themselves from COVID-19.

90% adopted at least one of these measures to protect themselves and others from COVID-19.

- 78% went into quarantine (voluntary or mandatory preventive isolation).
- 68% adopted over three hygiene and self-care measures.
- 60% adopted over three physical distancing measures.
- People aged 40 and over adopted higher percentages of hygiene and self-care practices to prevent COVID-19, while younger people (18-24 years old) adopted higher percentages of preventive behaviours associated with physical distancing.
- The greatest difference between women’s and men’s behaviours was the increased cleaning frequency at home or in the workplace, which was 52% among men and 62% among women.
- Cali (72%) and Bogotá (70%) adopted higher percentages of hygiene and self-care practices (Figure 2). Cartagena (65%) and Medellin (60%) adopted greater percentages of physical distancing practices. The municipalities with lower virus spread adopted lower percentages of hygiene and self-care practices (63%) and physical distancing (50%).

**Figure 2.**
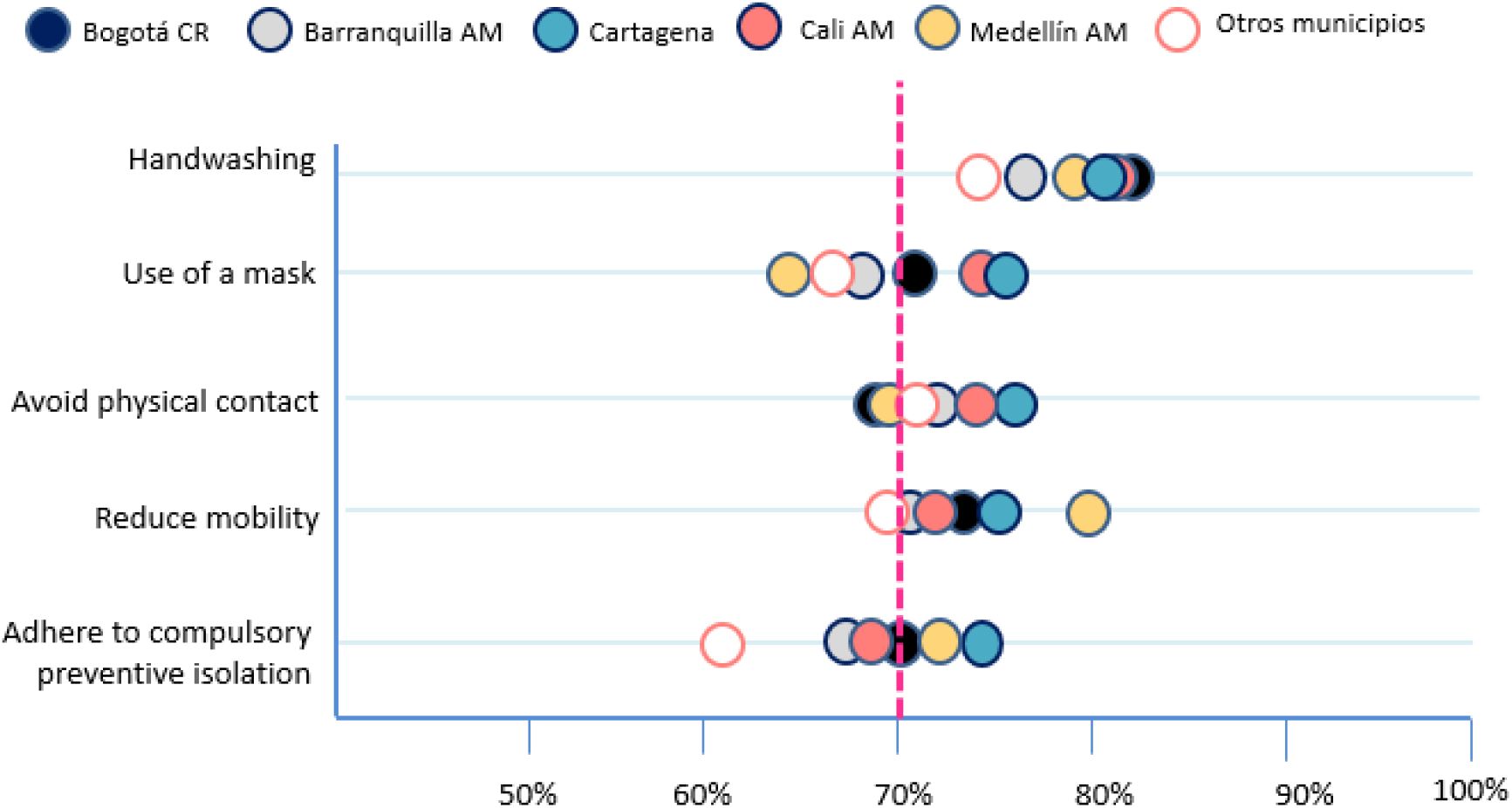
Compliance with Covid-19 control measures in Colombia in cities with high virus spread.

### 2. Self-isolation (voluntary quarantine)

46% voluntarily went into preventive isolation before government measures.

- No differences were found by sex.
- 54% of the 45-49-year-old group, 50% of the young group (18-24 years) and 50% of the older group (60 years and over) went into voluntary quarantine.
- 5% decided to isolate themselves because they experienced flu symptoms, fever, or cough.
- Only 2% adopted voluntary quarantine because they experienced flu symptoms, fever, or cough, and the local hospital or healthcare providers instructed them to do so.

Considering all the adopted isolation and physical distancing measures, the reasons for these behavioural changes were:

- 73% feel it is the right thing to do and contributes to containing the pandemic.
- 59% followed the national government’s guidelines for mandatory preventive isolation.
- 55% responded to the increase in cases and the collapse of health systems in other countries.
- 48% reacted to the increase in reported cases of COVID-19 in Colombia.
- 30% complied with drills, curfews and local, district, or territorial government measures.
- 30% responded to the closure of schools, colleges, and universities
- 16% complied with their boss’s orders.
- 13% complied with the recommendations of their healthcare provider or local hospital.
- 4% followed the recommendation of a friend or family member.

78% of the respondents voluntarily isolated or followed the government’s measure; only 33% started working from home. The percentage of people who started working from home is higher among the public (55%) and private sector employees (48%) and lower among the self-employed (28%), women heads of household (28%) and people affected by armed conflict (18%), indicating that conditions of informal workers and vulnerable populations are less flexible (Figure 3).

**Figure 3.**
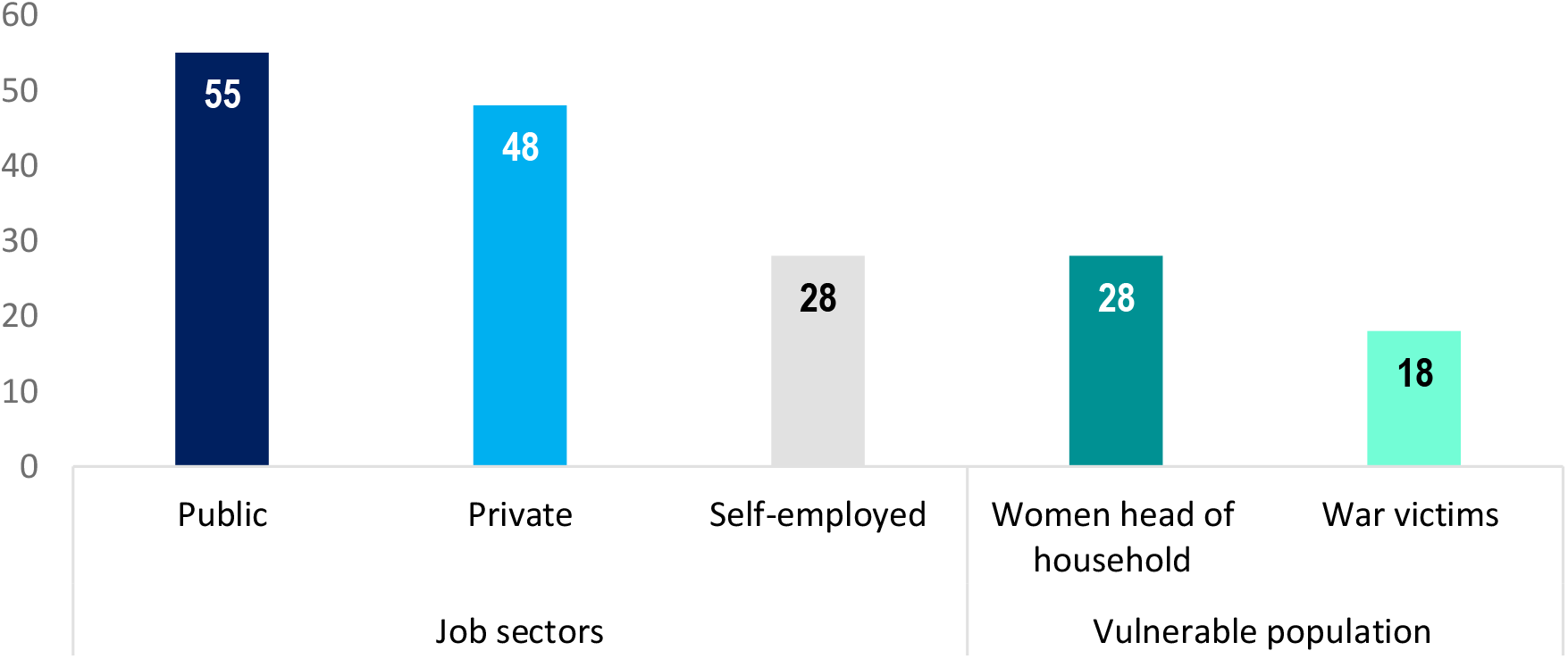
Percentage of people who started working from home by employment sector and vulnerable populations.

People over 59 years of age (85%) and people between the ages of 45 and 49 years of age (82%) adopted self-isolation (voluntary or mandatory) in higher percentages.

- 78% of women, 76% of men, and 70% of non-normative gender respondents adopted self-isolation (voluntary or mandatory).
- 78% of those who have a paid job, 77% of those who do not work and 74% of those who work without pay adopted the measures.
- 72% of people with a monthly family income of below 500,000 COP (apx. 130 US) entered voluntary quarantine; this percentage reaches 84% among people with a family income of over 10 million COP (apx. 2,100 US).
- Among those who adopted self-isolation, 11% of people with a monthly family income below 500,000 pesos have got savings; this percentage rises along family income and reaches 77% among people with a family income of more than 10 million COP.

Concerns related to COVID-19 and self-isolation among the respondents are:

- 79% infection of a family member.
- 74% lack of healthcare for a family member.
- 71% effects of the pandemic in the poorest and most vulnerable people.
- 69% failure to comply with government measures.
- 64% economic future and recession.
- 61% uncertainty about returning to normal life.
- 53% availability of a new vaccine or drug.
- 52% shortage of food, medicine, and medical supplies.
- 50% separation from other family members living alone.
- 44% inability to pay rent or utilities.
- 41% loss of job, income, and savings.
- 27% Anxiety and depression during isolation.
- 22% future education of their children.
- 20% being alone and unable to care for themselves.
- 8% not having a computer or access to the internet.
- 7% impact on children’s behaviour.
- 6% domestic violence.
- 4% effects on their social lives.

### 3. Groups that react under quarantine

Preliminary results using the *k-means* technique allowed us to identify three groups that are reacting differently to the pandemic in the way they are changing behaviours, and facing physical distancing measures (4):

- those who accept it (40%)
- those who resist the situation (34%)
- those who suffer because of it (26%).

Young people are more likely to be among those who resist and suffer from the situation, while those over the age of 49 are in the group of those who are most accepting of the situation. Men are more likely to resist, and women are having a harder time than men: almost two-thirds are in the group of those who are suffering during the current situation. Women over 39 are also much more likely to accept quarantine, whereas younger women are much more likely to suffer as a result (Figure 4).

Some characteristics of each group are highlighted below (Figure 5).

**Figure 4.**
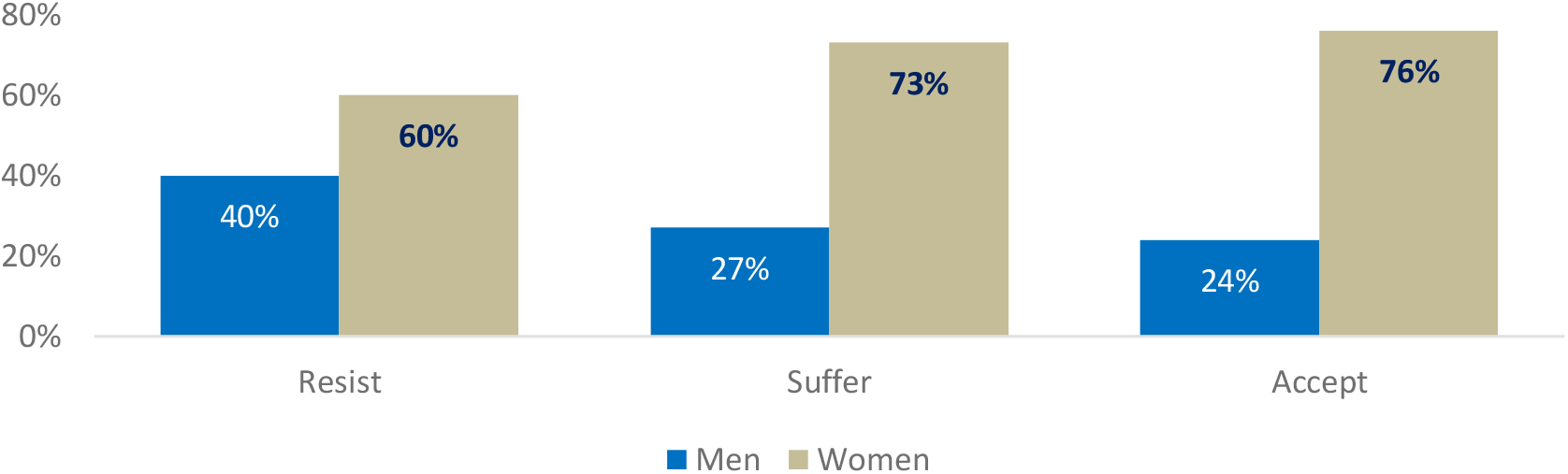
Reactions to quarantine measures by gender in Colombia.

**Figure 5.**
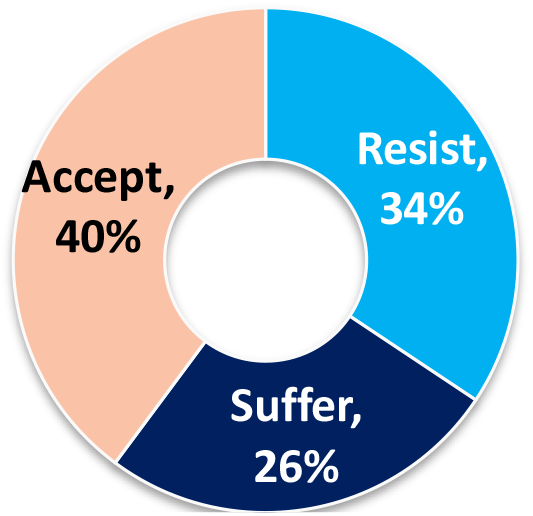
Groups that react under quarantine in Colombia.

### Group 1. Resist

In the group which resists, 40% are men, 68% are under 39 years old, 40% have savings, and 63% have not had any chronic diseases. In response to COVID-19, this group is characterized by the following:

64% adopted less than four physical distancing measures. In the lowest percentages:

- they adopted hygiene measures.
- they avoided going out to social events or using public transport.
- they avoided contact with people who have a fever or respiratory symptoms.
- they avoided contact with people who had travelled in the last month.
- 12% have shown signs of nervousness, anxiety, or depression.

More than 85% expressed they had no worries associated with domestic violence or living with others.

### Group 2. Suffer

In the group who is suffering from the pandemic, 73% are women, 64% are under 29 years old, 55% have an average family income over 2 million COP (apx. 511 US), 61% have had some chronic illness or somebody in the family. In response to COVID-19, this group is characterized by the following:

- 68% adopted more than three measures of physical distancing.
- 73% have shown signs of nervousness, anxiety, or depression.
- Over 80% are anxious because:
  - someone in the family may get infected with COVID-19.
  - someone in the family may have a medical emergency and not receive care.
  - the pandemic will not be under control and uncertainty on how or when normal life can resume.
  - Economic future and recession.
- In smaller percentages, they have been able to do daily exercise or have tried to eat healthy.

### Group 3. Accept

In the group of people who adapt to the situation, 76% are women, 43% are over 49 years old, 36% have savings, and 63% have not had chronic illnesses. In response to COVID- 19, this group is characterized by the following:

- 63% adopted more than three measures of social distancing
- 73% have shown signs of nervousness, anxiety or depression
- They do the following in higher percentages:
  - communicate with family and friends
  - use social media to stay informed and connected
  - do exercise at home
  - cook
  - try to eat healthy
- 83% believe that by complying with mandatory social isolation, they are contributing to stopping the pandemic.
- 60% consider that quarantine has allowed them to reflect on global, collective, and personal health issues.

In most cases - 63% and 68% - all three groups supported physical distancing measures. Those who resist show less support for physical distancing measures (Figure 6).

**Figure 6.**
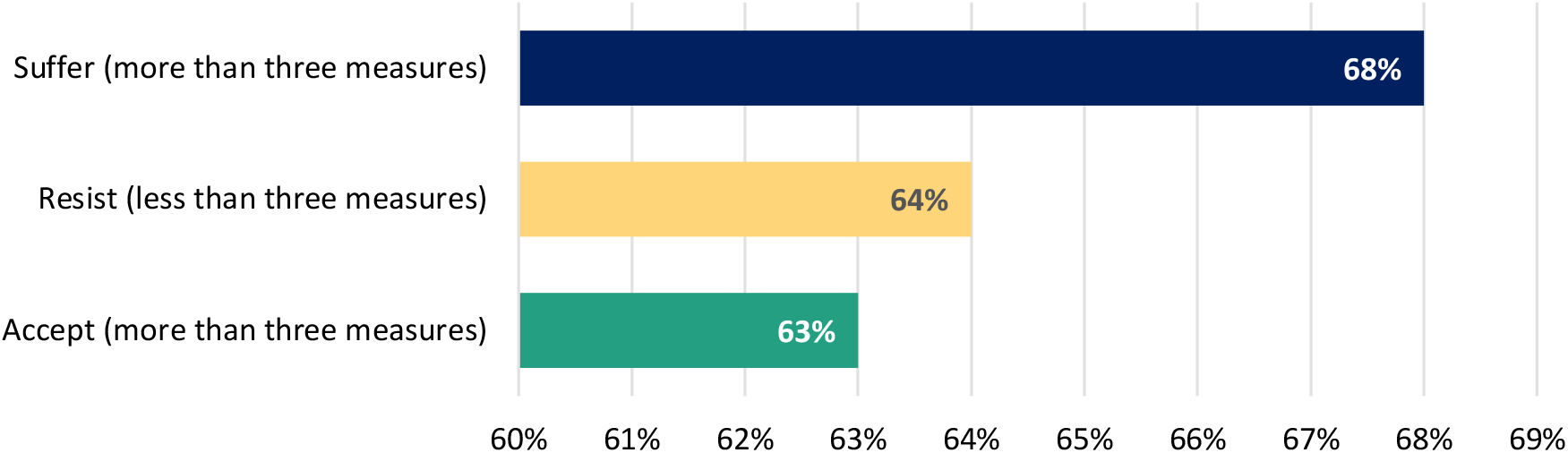
Compliance with physical distancing measures by type of reaction in Colombia.

### 4. Social response and protective measures in Colombia, Hong Kong, and the UK

The social response to government pandemic control actions can be measured by how people have adapted their behaviours to protect themselves and others from infection. Considering the experience of some countries that implemented interventions to reduce the demand for health services and mortality. Below is a comparison of three countries that conducted the survey (Figure 7), highlighting five specific measures to control COVID-19 in each country. From analysis, the following can be highlighted:

**Figure 7.**
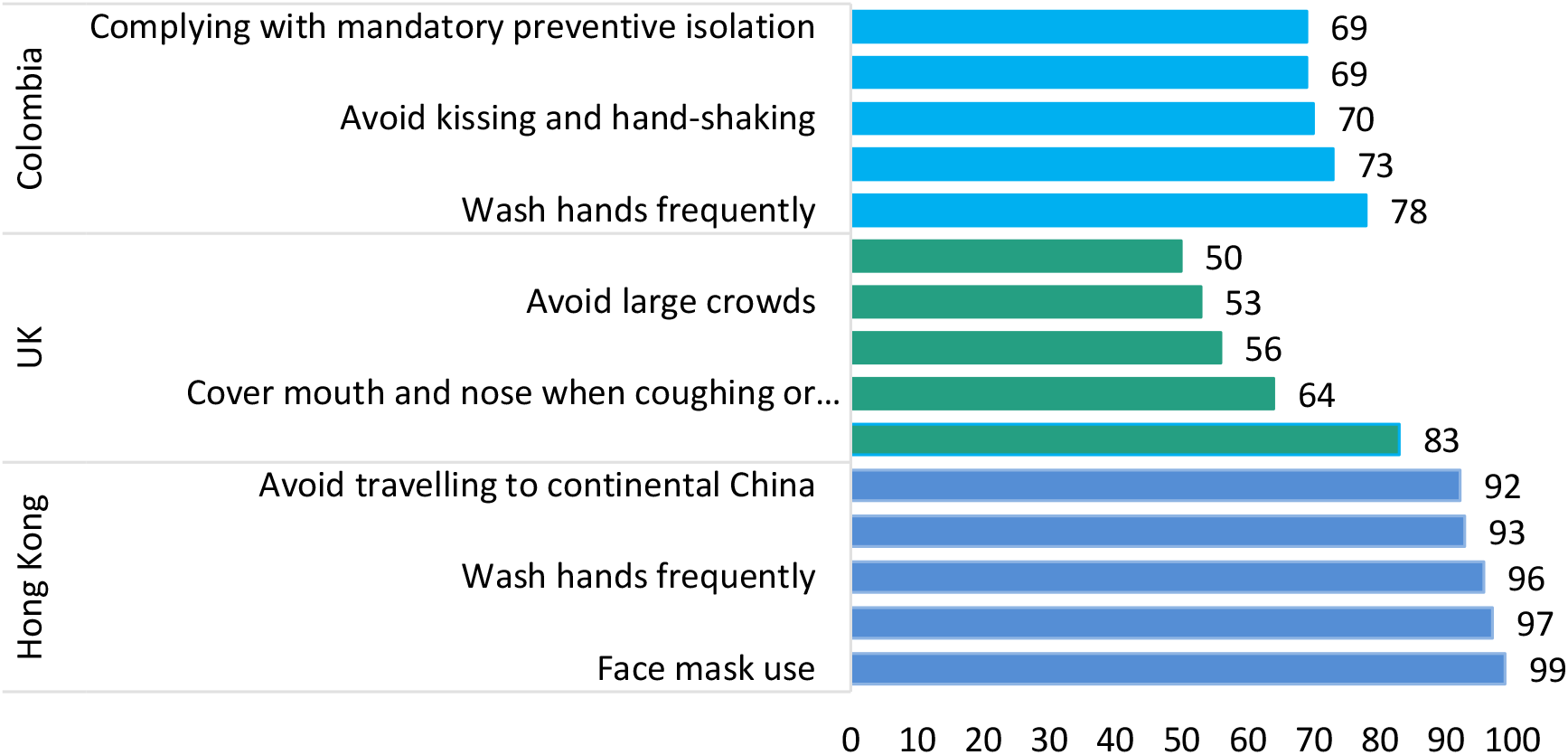
Most frequently adopted COVID-19 protective measures in Hong Kong, the UK and Colombia.

- In Hong Kong, compliance levels with measures and behavioural changes are above 90%. In the UK, compliance has greater variability (ranging from 83% to 50%), and in Colombia, these levels are between 78% and 69%.
- Hand-washing is frequent in all three countries: Hong Kong (96%), the UK (83%), and Colombia (78%), while in Hong Kong this is the third most common practice, in the UK and Colombia it is the most common.
- In Hong Kong, most of the population (99%) adopted the use of face masks; in Colombia, this practice was adopted by 69% of the people and ranked fourth.
- Covering up when coughing or sneezing (97% in Hong Kong) and covered sneezing (64% in the UK) rank second among these countries and in Colombia, it is not ranked among the first five adopted preventive behaviours.
- In Hong Kong, 93% of the people avoid contact with people with respiratory disease symptoms; the UK (about 50%) and Colombia (about 70%) adopt physical distancing measures such as avoiding going out or attending social events.

### 5. Care and employment responsibility

50% is responsible for the care of a person in their household.

- 22% are responsible for the care of children between the ages of 5 and 17.
- 16% are responsible for the care of people over the age of 65.
- 15% are responsible for the care of individuals between the ages of 18 and 64.
- 11% are responsible for the care of children under 5 years of age.
- 2% are responsible for the care of a person with a disability.

52% of women and 45% of men are responsible for the care of a person in their household.

57% of residents in male-headed households report an average family income of more than 2 million COP (apx. 511 US); 68% have no savings; 38% started teleworking; 22% are unemployed.

76% adopted isolation (voluntary or mandatory); 26% left home during quarantine to buy medicines, 12% to help a dependant, and 9% to go to the hospital.

16% of women are heads of household, and among them, 83% are responsible for the care of a person in their home.

- Among women heads of household who have a dependent, 57% have an average family income of less than 2 million COP; 85% have no savings; 30% started teleworking, 28% are unemployed.
- 74% adopted isolation (voluntary and compulsory); 24% left home during quarantine to buy medicine, 11% left home to help a dependant, and 12% went to a hospital.
- 22% are in charge of children under 5 years old, 52% are in charge of children between 5 and 17 years old; 32% are in charge of people between 18 and 64 years old; 33% are in charge of people over 64 years old, and 6% are in charge of people with disabilities.
- The highest percentages of women heads of household with dependents can be found in Bogotá and its surrounding municipalities (33%), Cali and its metropolitan area (16%), and other municipalities with lower virus spread (29%).

### 6. Risk perception, unmet sexual, reproductive and mental health needs during the pandemic

98% consider that COVID-19 is a severe health problem; 80% believe that COVID-19 can cause serious health consequences for the whole population without exception; 17% think that COVID-19 can cause serious health consequences for some risk groups such as adults over the age of 65.

People feel that it is very likely or likely to become infected with COVID-19 by using public transportation (88%); having contact with someone infected (87%); having contact with contaminated surfaces (86%) and having contact with the soles of shoes, sneakers or flip-flops (69%) more than by consuming or using products imported from China (21%), eating meat from wild animals (21%) and living with a dog or cat (6%) (Figure 8).

**Figure 8.**
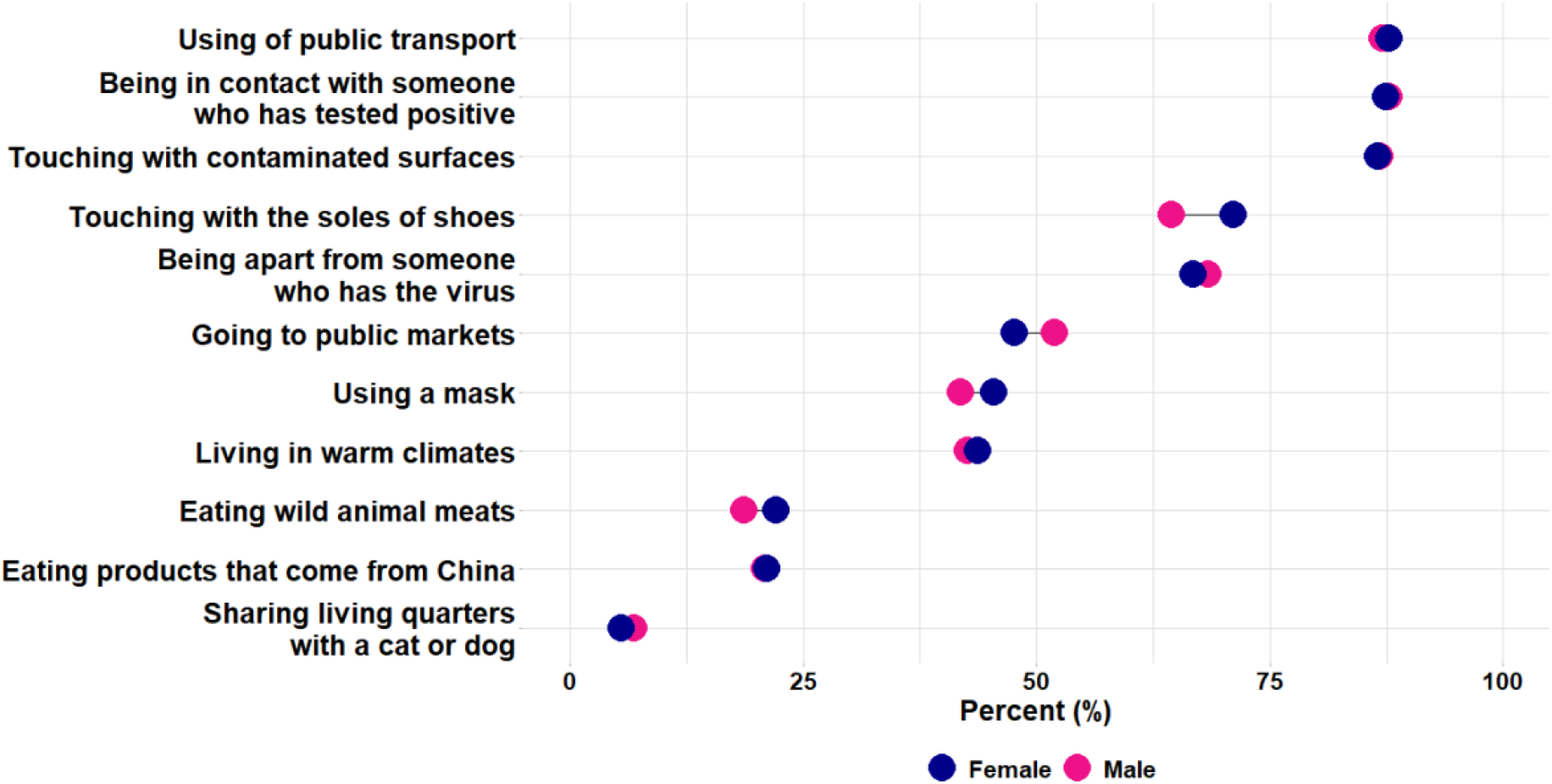
Percentage of people who believe it is likely, or highly likely to become infected with COVID-19 according to the following situations (by sex):

People feel that it is likely or highly likely to prevent the spread of COVID-19 by adopting the following practices:

- 21% - drinking water every 15 minutes, fluids, or hot drinks.
- 18% - gargle (throat wash) with baking soda.
- 19% - cleaning the house with eucalyptus smoke.
- 12% - consuming garlic.

Figure 9 shows the following:

- The highest percentage of people over 50 years of age who believe it is likely or highly likely that COVID-19 is transmitted through contact with contaminated surfaces is in Barranquilla.
- The lowest percentage of young people (18 to 29 years old) with the most significant variation in risk perception from transmission from one city to another was found in Cartagena.
- Barranquilla has the highest percentage of people between the ages of 30-39-year-olds, and Cali the lowest. The 40-49 age group presents the highest percentage of those who think it is likely or highly likely that COVID-19 will be transmitted between the municipalities with the lowest levels of virus spread is higher. In Cartagena, this percentage is again lower.

Figure 10 shows that:

- Among young people (18 to 29 years old), the percentage that considers the transmission of COVID-19 using public transport to be probable or highly probable is higher in Cartagena and lower in Cali.
- Among those aged 30 to 39, with lower variation, the highest percentage is also found in Cartagena, the lowest in Barranquilla. In the 40-49 age group, Cartagena stands out with the lowest percentage of people who consider the spread of COVID-19 through public transportation to be likely or highly likely, whereas Medellin has the highest.
- Among the oldest group (50 years and over), we found the most significant variation between municipalities, with Barranquilla having the lowest percentage and Cartagena the highest.
- Regarding health perception: 76% consider their health condition to be good or very good.
- 97% did not take the test, and 2% preferred not to answer the question; 1% took the test (30 cases): 0.48% took the test and the result was negative (17 cases); 0.34% took the test and have not yet received the result (12 cases), and 0.03 took the test and their result came back positive (1 case).
- 43% mention any of the diseases included in the survey; among the most frequent are:
- 17% ear, nose, or throat problems.
- 10% mental illness.
- 7% chronic diseases.
- 6% cardiovascular diseases.
- 5% intestinal problems.
- 5% respiratory diseases.

Regarding unmet sexual and reproductive health (SRH) needs in the context of COVID-19:

- 20% of people have had some need for sexual and reproductive health (SRH) care.
- Young people (18 to 29), people with family incomes of less than 500 thousand pesos and Venezuelan migrants have higher percentages of unmet sexual and reproductive health needs, exceeding 25%.

Among women, reproductive health needs and the reasons why they have not been met are:

- 21% did not access these services because they preferred to stay in quarantine.
- 13% of women need access to contraceptive methods.
- 12% of women require a gynaecological visit.
- 6% did not have access because their healthcare provider suspended their services.
- 4% did not have access to these services because they could not afford them.
- 3% did not have access because there is no telehealth service available.

Amongst men, reproductive health needs and the reasons why they have not been met are:

- 17% did not access these services because they preferred to stay in quarantine.
- 3% of men require a urology consult and need to take STD tests, respectively.
- 3% did not have access to these services because they could not afford them.
- 2% of men need access to contraceptives.
- 2% did not have access because their healthcare provider suspended their services.
- 2% because there is no telemedicine service available.

75% have experienced issues with their mental health in the last 21 days as a result of current preventive measures: 54% felt nervous; 52% have felt tired for no reason; 46% felt restless and impatient, and 34% felt anger and rage (Figure 11).

Figure 12 highlights the most common mental health issues during the last 21 days:

- The 18-29-year-old group felt more nervous (61%), tired for no reason (62%), impatient (60%) and angry (48%) during the past 21 days, compared to the age group over 50 who felt less nervous, tired for no reason, impatient or angry.
- Among the 50 and older age group, feeling nervous (41%) was more common during the last 21 days and perhaps the most highly reported mental health issue.

Nearly three out of four people (73%) who are suffering and who are accepting the situation report being more anxious and depressed than usual. Only one in ten (12%) people resisting report issues on their mental health (Figure 13).

**Figure 9.**
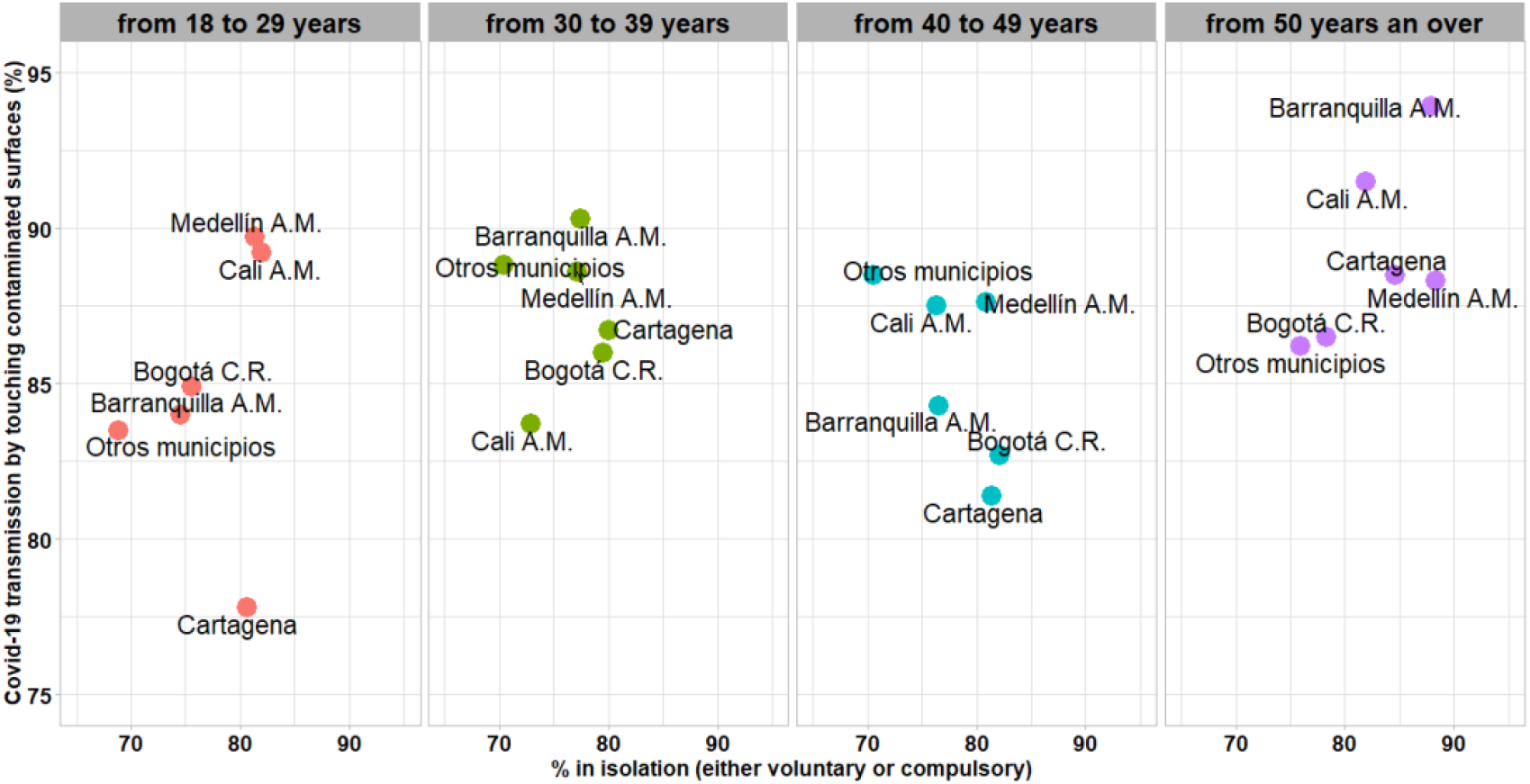
Percentage of people who went into voluntary quarantine and consider it likely or highly likely to contract COVID-19 through contact with surfaces by cities and age groups in Colombia.

**Figure 10.**
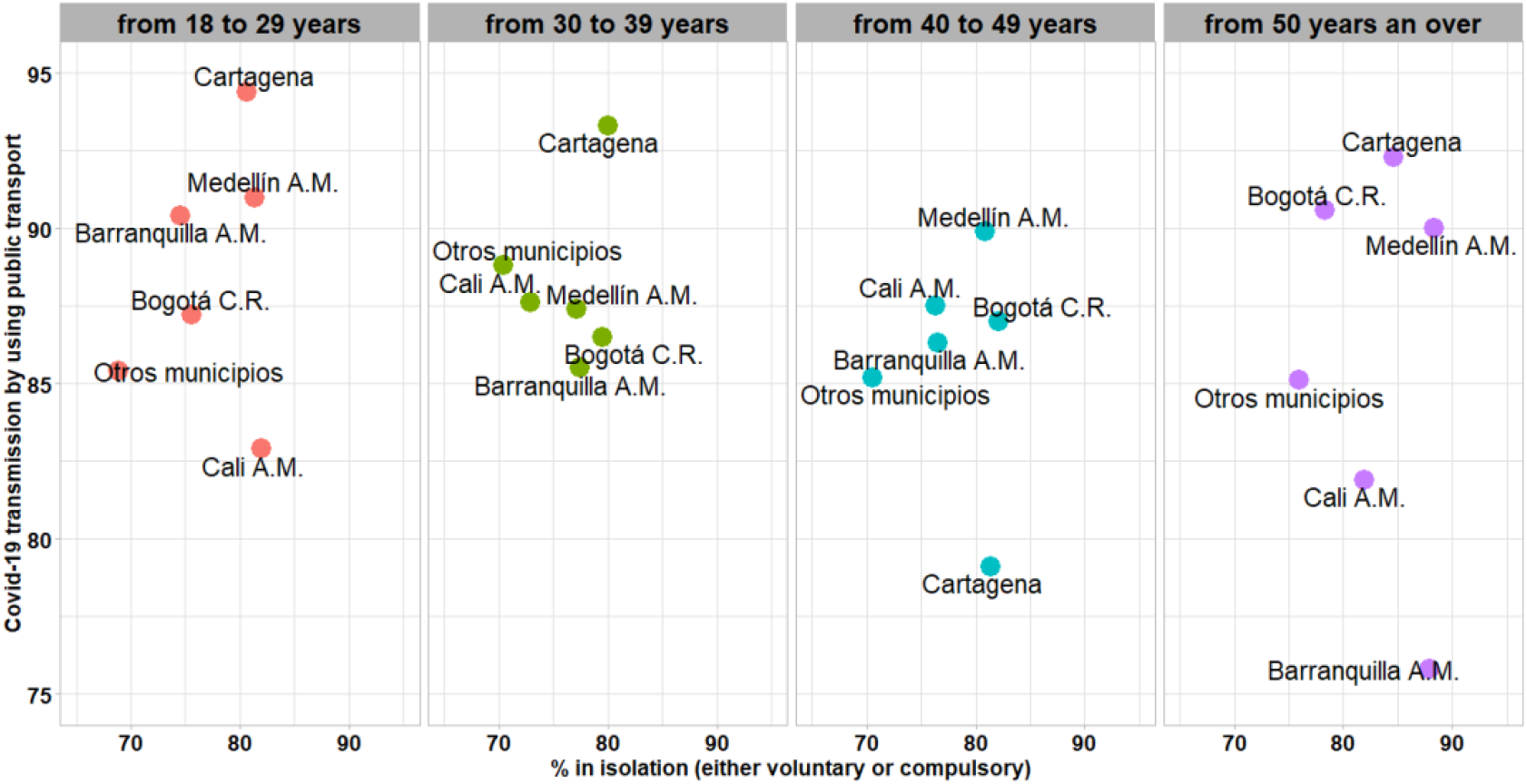
Percentage of people in isolation who consider COVID-19 transmission by using public transportation to be likely or highly likely, by municipalities and age groups in Colombia.

**Figure 11.**
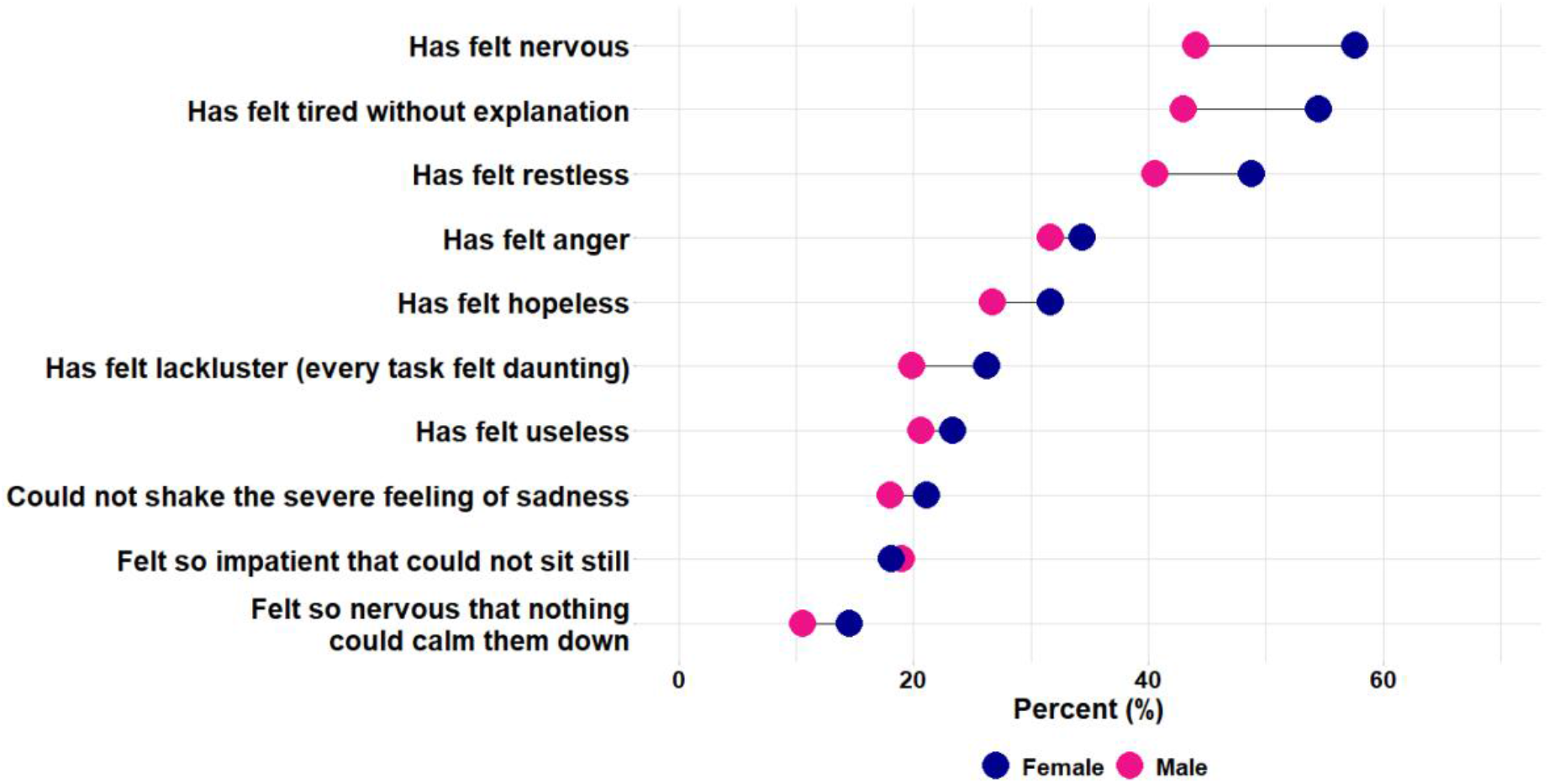
Percentage by sex of people who have felt any of the following mental issues in the last 21 days.

**Figure 12.**
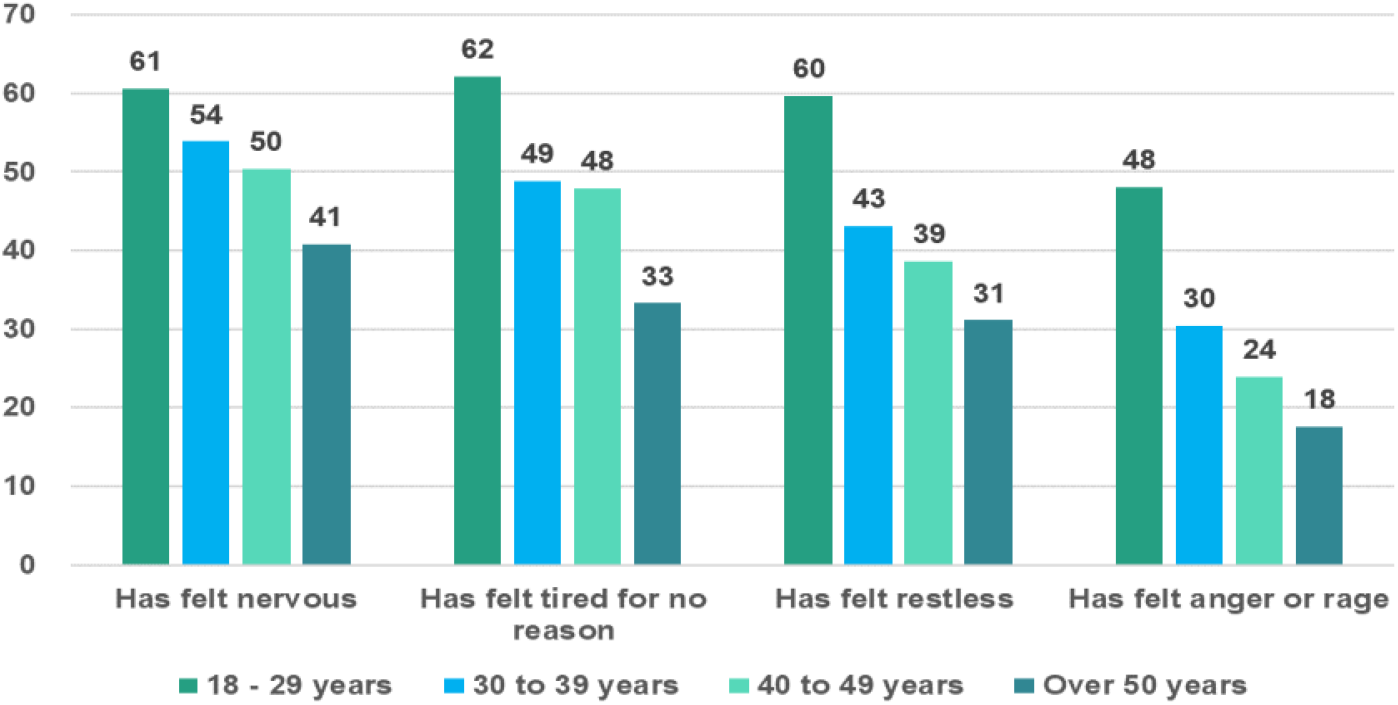
Percentage of people who have felt any of the following mental issues in the last 21 days, by age group in Colombia

**Figure 13.**
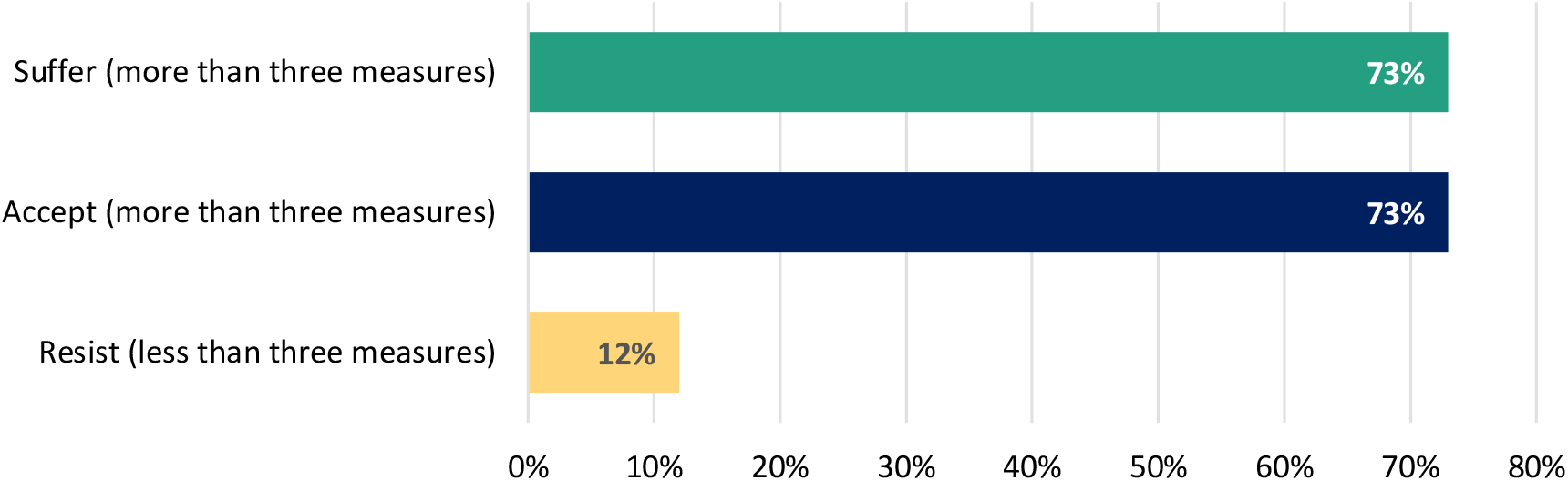
Percentage of people saying they feel more anxious and depressed since the announcement of the isolation measures by quarantine reaction groups.

### 7. Access to quality information

People are getting information about COVID-19 from the following sources:

- 63% official websites
- 58% social networks
- 54% television
- 35% communications from work, school or university
- 31% family and friends
- 28% newspaper/magazines (print or digital)
- 20% the radio
- 9% other support networks
- 6% doctor or other healthcare professionals
- 3% religious communities.

The top three most popular media are the same in both genders; no significant differences were found.

- Access to information on COVID-19 through television increases with age (from 52% under 25 years to 70% over 59 years).
- Access through official websites is high among young people and presents the highest value in the 35-39 age group (69%) but decreases to 52% among older people.
- Access to information on COVID-19 through social networks has less variation, being higher among the youngest (64%) and close to 53% from age forty onwards.
- People who have not adopted isolation (voluntary or compulsory) have the lowest percentages of access to information on COVID-19 through any media source. Among the most used media, access through official websites reaches 70% among those who adopted isolation and drops to 35% among those who did not; access through social networks drops from 65% to 36%, and television access drops from 60% to 35%.
- Cali and Medellin have the highest percentage of COVID-19 information access through official websites (67%) and the lowest access through television (46% and 51% respectively). Social media access does not vary considerably between municipalities.
- 1% do not receive any information on COVID-19. This percentage reaches 2% in Barranquilla and in the municipalities with low spread of the virus.

### 8) Government response perception

In Colombia (Figure 14):

**Figure 14.**
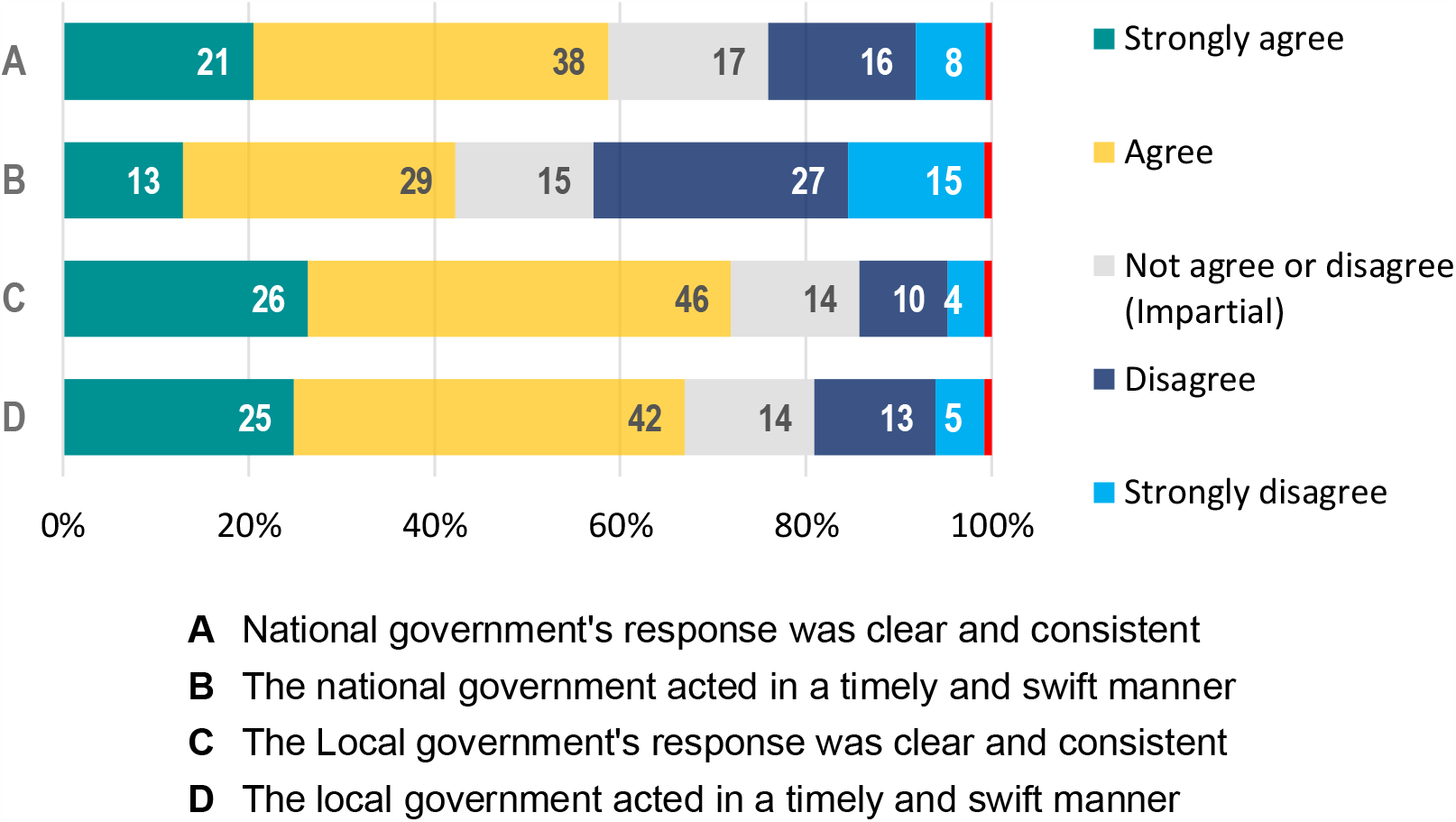
Perception of national and local government response to control COVID-19 in Colombia.

- 59% agree or strongly agree that the national government’s COVID-19 control response was clear and consistent.
- 42% agree or strongly agree that the national government acted promptly to control the spread of COVID-19.
- 72% agree or strongly agree that the local government’s COVID-19 control response was clear and consistent.
- 67% agree or strongly agree that the local government acted on time to control the spread of COVID-19.
- Women have higher percentages, a favourable perception (strongly agree or agree) with the level of response and timeliness shown by national and local governments.
- Young people (18-30 years old) have the lowest favourable perception of the level of response and timeliness shown by national and local governments.
- Bogota shows the highest favourability regarding the level of response (82%) and timeliness by local authorities (78%). Cartagena follows with 74% and 75%, respectively.
- People who had a favourable perception of national and local government response to control the transmission of COVID-19 adopted preventive isolation measures (voluntary and mandatory) in percentages over 85%.

## Data Availability

This database is freely accessible and for research purposes, if you use it, we would like to acknowledge in your work and material obtained the credit and authority of the collection, storage and purification of the Profamilia Association.

https://www.profamilia.org.co/wp-content/uploads/2020/05/Base-de-datos-encuesta-Solidaridad.sav

https://profamilia.org.co/investigaciones/estudio-solidaridad/

## Acknowledgements

This study was funded by Asociación Profamilia.

